# Contribution to COVID-19 spread modelling: A physical phenomenological dissipative formalism

**DOI:** 10.1101/2020.05.02.20088971

**Authors:** Oualid Limam, Mohamed Limam

**Affiliations:** University of Tunis El Manar, Ecole Nationale d’ingénieurs de Tunis, Laboratoire de génie civil; Medical Doctor (immunologist), laboratory of medical biology, Regional Hospital of Sidi Bouzid, Tunisia

**Keywords:** COVID-19, Transmission, lattice, wave, free energy, dissipation

## Abstract

In this study, we propose an evolution law of COVID-19 transmission based first on a representation of population by a domain part of an infinite ordered lattice in which epidemic evolution is represented by a wave like free spread starting from a first case as an epicentre. Free energy of spread on a given day is defined equal to the natural logarithm of the number of infected cases. Dissipation of propagation is obtained using a postulated form of free energy built using thermodynamics of irreversible processes in analogy to isotherm wave propagation in solids and elastic damage behaviour of materials. The proposed expression of daily free energy rate leads to dissipation of propagation introducing a parameter quantifying measures taking by governments to restrict transmission. Entropy daily rate representing disorder produced in the initial system is also explicitly defined. In this context, a simple law of evolution of infected cases as function of time is given in an iterative form. The model predicts different effects on peak of infected cases Imax and epidemic period, including effects of population size N, effects of measures taking to restrict spread, effects of population density and effect of a parameter T similar to absolute temperature in thermodynamics. Different effects are presented first. The model is than applied to epidemic spread in Tunisia and compared with data registered since the report of the first confirmed case on Mar 2, 2020. It is shown that the low epidemic size in Tunisia is essentially due to a low population density and relatively strict restriction measures including lockdown and quarantine.

## 1. Introduction

First cases of pneumonia unknown etiology have been declared in Wuhan, China since Dec 8, 2019. Pneumonia starts with severe acute respiratory infection symptoms and some cases developed acute respiratory distress syndrome with failure complications. On Jan 7, 2020, Chinese centre for disease control and prevention identified a new coronavirus (Chen et al, (2020)).

COVID-19 is a human coronavirus include in the gender beta coroanvirus group 2b, family coronaviridae. It is the third strain of virus of the coronavirus family (CoV), isolated in humans in the context of an epidemic after SARS-CoV in China (2002) and MERS-CoV in Saudi Arabia (2012). Examination of the COVID-19 genome showed genetic similarity to SARS-CoV about 79.5%. Human to human transmission takes place by either respiratory droplets or close contacts. According to the world health organization, COVID-19 is a unique virus that causes respiratory disease and which spreads via oral and nasal droplets (Kolifarhood et al, 2020).

On Mar 2, the first case has been declared in Tunisia.

Actually, COVID-19 is causing a disease representing a planetary problem for public health and negative impact on humanity (Boccaletti S. et al, 2020).

The objective of this paper is to propose a simple model to predict COVID-19 transmission using early data of the outbreak. Majority of epidemic transmission models are based on mathematical approaches dividing population in different interacting groups and assuming different rates of transmission between them. Solutions are conducted using integration of differential equations and principle of conservation (Kermack, W.O. and MC Kendrick, A.G., 1927). Population is generally assumed as a closed system, the probabilistic formalism of transmission between individuals of different groups leads to a population size effect on epidemic size and epidemic period. Models that are more sophisticated include also Monte Carlo numerical simulations for stochastic models and more realistic epidemic networks. See for example a review by House et al, (2013) and recent studies by Kim et al, (2020), Liang, (2020), Li et al (2020) among several others for COVID-19 modelling.

Epidemic networks and lattice methods have their origin in social science and computer science (See for example a review by Keeling and Eames (2005)). Lattice models are representation of an ordered network in which epidemic transmission is similar to a wave like spread in regular grid representing connected individuals. Epidemic starts from an epicentre and spreads out in a roughly circular manner. Figure 1 illustrates an example in two dimensions. Lattice models are suitable for example for forest-fire models (Bak et al, 1990) where nodes represent trees that burn leaving empty sites. Keeling and Eames (2005) interpreted this representation as similar to epidemic transmission.

**Figure 1.**
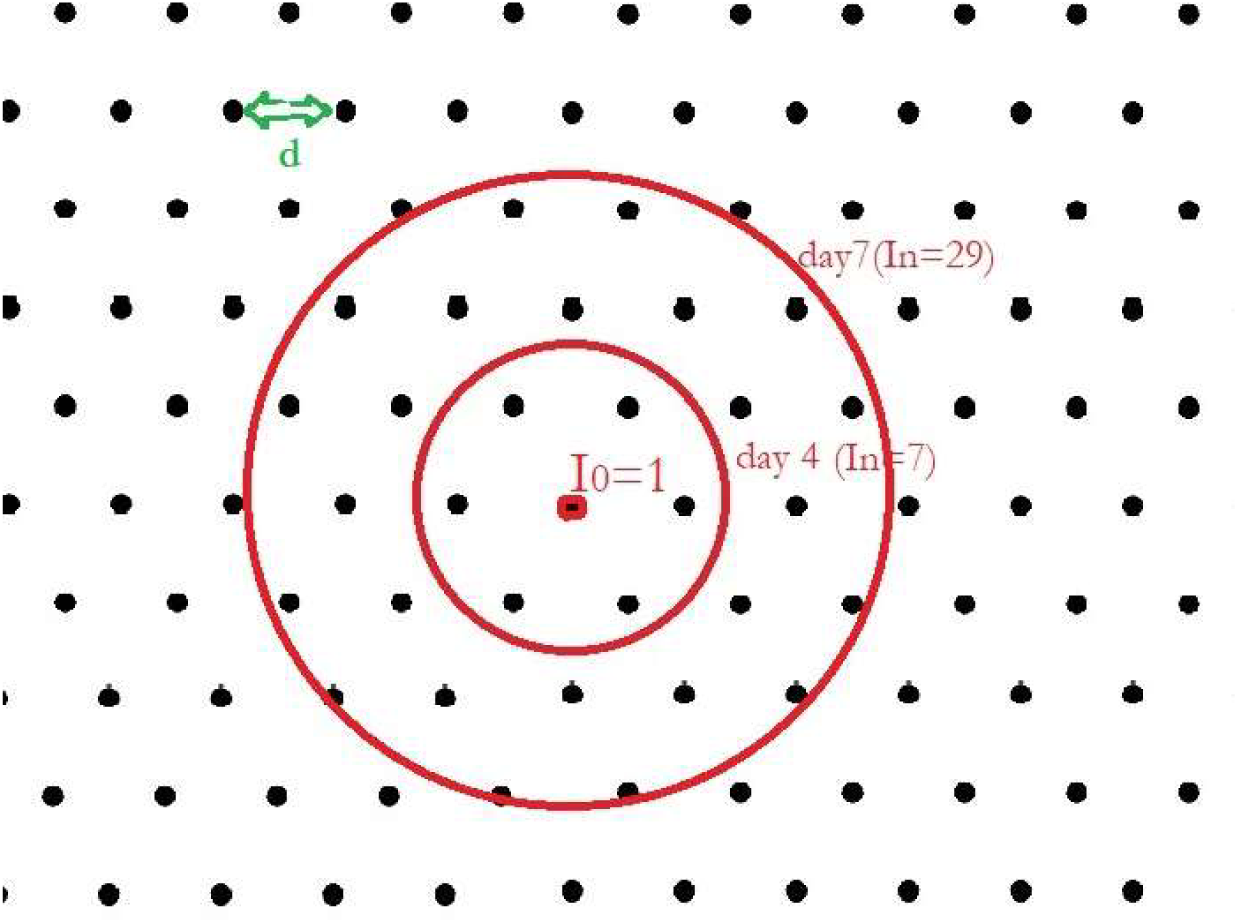
Lattice representation of population with a wave like epidemic spread.

This paper proposes, in this context of lattice representation, a law of propagation and dissipation using a formalism in analogy to elastic wave propagation and a size and temperature dependent elastic damage material model ((Ben Hassine et al (2019) and (Limam et al, 2014).) The advantage of a formalism inspired from thermodynamics is that different effects emerge from principles.

## 2. Materials and Methods

The proposed model is first presented and explained in Section 3. In Section 4, the model is applied to study different effects including measures to restrict spread, effect of population density and size and effect of a parameter T similar to absolute temperature in thermodynamics. Epidemic evolution in Tunisia is also analysed. We consider for comparison, data from national observer for new and emergent diseases (http://www.onmne.tn) until Apr 25.

## 3. Theory

Consider first a simple mathematical model given by Equation (1) and (2). *I_n_* is the number of infected people on day *n*. Theses Equations correspond to epidemic theoretical free transmission in a population of size N defined in a finite convex domain part of a perfect infinite ordered lattice of connected people and starting from an epicentre *I*_0_ = 1 belonging to the domain. Population density is inversely proportional to the square of distance d depicted in figure 1. Coefficient C defined by Equation (1) is considered as an intrinsic characteristic of population density and independent of population size. It is clear that it decreases when distance d increases which means that when population density decreases. It represents the number of transmission between every infected person at wave front to other persons. A theoretical free transmission in the lattice corresponds to the linear curve with a slope ln[C] in a semi-logarithmic scale depicted in figure 2 (a, b) for a domain representing, for example, a typical dense city with N=12 Million and C=1.62. In that case population size N will be reached at 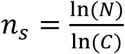, on day 34, meanwhile epidemic will continue to propagate in the lattice outside the population domain, as the considered population is fixed but transmission to the outside was made possible by hypotheses.

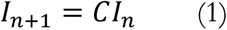

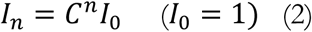

**Figure 2.**
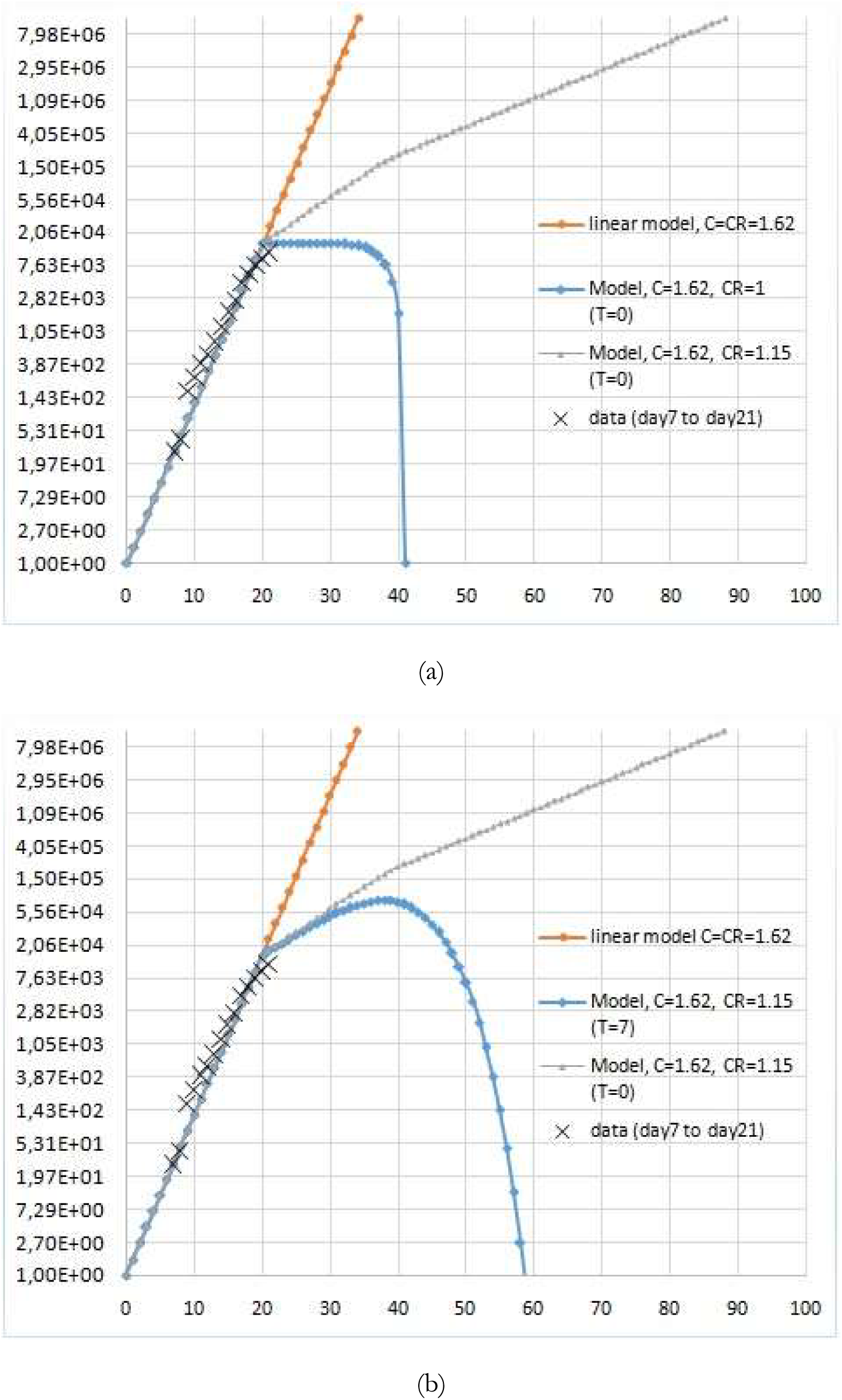
Infected cases as function of time (day n): Analogy with wave spread in elastic damage material

In reality transmission process is dissipative and the linear curve of slope ln[C] corresponds only to early stage. C can be deduced, for example, from initial data curves fitting in a semi-logarithmic curve as shown in figure 2. This coefficient should be reduced by measures imposed by governments including lockdown and quarantine, or by population behaviour including social distancing, personal hygiene, for example by wearing a mask in fear of the spread of the virus similarly to the effect described by for example by Kim et al, (2020) or Liang, (2020). This behaviour emerges naturally after first deaths inducing a disorder in the initial lattice of figure 1, which means physically that entropy should increase.

The idea of the proposed model can be highlighted when we made an empirical analogy of the linear curve depicted in figure 2 with energy as function of time of an elastic wave propagation in a rod obeying Hooke’s law of elasticity and submitted to harmonic imposed power. So we defined by analogy free energy of the virus spread by *φ* = ln(*I_n_*) which gives a constant daily rate in the case of linear curve depicted in figure 2 and defined by Equations (1) and (2).

In reality, material behaviour as epidemic spread is dissipative and wave velocity will decrease due to material damage. In analogy to reduction of Young modulus of elasticity in damage mechanics (Kachanov, 1958), we should introduce a reduction of C in a semi-logarithmic scale. So we postulate the evolution model of infected cases *I_n_* on day n, given by Equations (3–7), where T is a parameter similar to absolute temperature in thermodynamics and where a first case is *I_0_ –* 1.

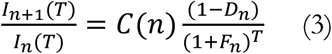

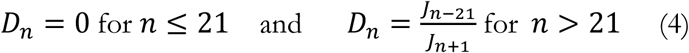

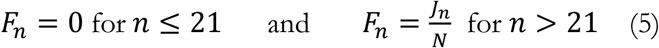

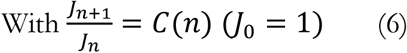

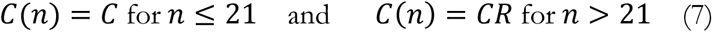

In order to give a physical sense to these Equations, we define first a free energy rate of the virus noted *φ_n_*_+1_ − *φ_n_ −* Δ*φ* given by Equation (8). In a thermodynamically consistent isotherm framework, Helmholtz free energy is defined by the rate Δ*φ* = Δ*U − T*Δ*S*, where Δ*U* is internal energy rate and Δ*S* is entropy rate given respectively by Equations (9) and (10) in the case of virus spread analogy and identified from Equation (8).

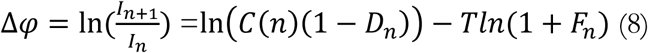

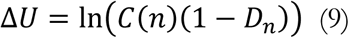

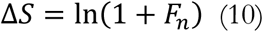

In order to consider lockdown and quarantine effect, C is decreased from day 21 and noted CR, a coefficient between 1 and C (Equation 7). Equation (4) is introduced to model recovering or death from day 21 with parameter *D_n_*. This is justified by recent studies reporting that observed duration of viral shedding among survivors was between 8 days and 37 days Zhou et al (2020). Evolution law of damage given by Equations (4, 6 and 7) is independent of population size and temperature and can be considered as an intrinsic property of virus spread. Its application with CR=1.15 ignoring entropy dissipative effect which means with T=0, leads to the trilinear curve presented in figure 2 (a and b) where population size will be reached with an epidemic period *n_s_* = 89 days for the considered example in Figure 2. Figure 2(a) shows also the theoretical case of an ideal lockdown with CR=1 and T=0 which leads to an epidemic period of 40 days due to a recovering rate higher than infection rate in that case with an epidemic peak of 11200, reached on day 21 and independent of N. Evolution law of damage given by Equations (4, 6 and 7) are defined exclusively by C and CR and remain independent of population size. This evolution law of damage induces a decreasing of internal energy rate defined by Equation (9) which remains also independent of population size and temperature and equal to free energy rate when T=0. Effects of T and N are rather due to entropy production. Similar hypothesis was considered for a damage evolution law as an intrinsic characteristic of the material independent of specimens size and temperature see (Ben Hassine et al (2019) and (Limam et al, 2014).

Equation (5) is introduced to consider population size effect, also from day 21. It is worth mentioning that the considered free energy is choosing with an entropy rate Δ*S* = *ln*(1 + *F_n_*), null before damage initiation (*F_n_* = 0 for (*n* ≤ 21)) and always positive, which means that entropy increases according to the second law of thermodynamics and contributes to dissipate free energy of the virus. Theoretical free transmission in an ordered lattice case given by Equations (1) and (2) can be obtained when considering CR=C, and T=0, in analogy to absolute zero state in thermodynamics, where entropy effect vanishes. Parameter T should be understood as for example hygiene measures in the system which can be linked also to ultraviolet rays increasing with ambient temperature rising. When increased it contributes to increase entropy effect and consequently to decrease free energy rate and epidemic spread. This is in agreement with recent environment studies shown also through statistical analysis of data that transmission decreases as ambient temperature increases, see for example ((Prata et al, 2020) and (Liu et al, 2020)).

Figure 2(b) shows an example applying the proposed model with N=12 Million, C=1.62, CR=1.15 and T=7. Introducing entropic effect, epidemic size is decreased with a peak of 79000 infected cases and an epidemic period of 56 days. Parameters were chosen to give an order of epidemic comparable to a dense city like Wuhan (Liang, (2020), Li et al (2020)). C was identified from the first slope of data using a regression analysis between day 8 and day 21.

## 4. Results

### 4.1. Restriction measures (CR) effect

Figure 3 presents an example of the model applied first with N=12Million, C=1.62, CR=1.15 and T=7 in a Cartesian scale and then with the same parameters but more restricted measures traduced by a reduction of C on day 21 to CR=1.1. It can be observed that CR=1.1 describes stricter measures that induce a decreasing of epidemic size and slightly increase epidemic period. The maximum of positive cases Imax is decreased from 79000 to 46000.

**Figure 3.**
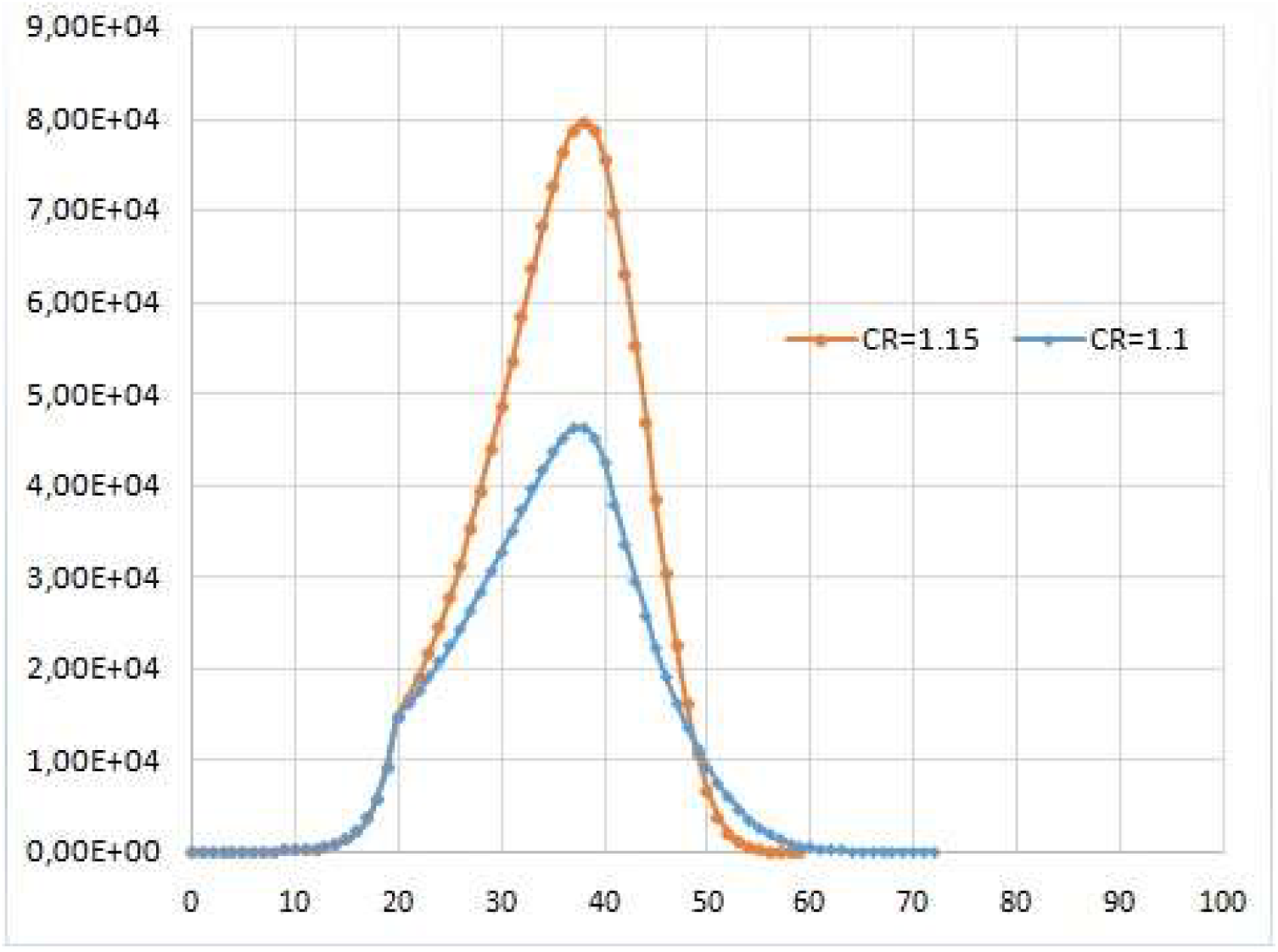
Restriction measures effect on infected cases curve (N=12E6, C=1.62, T=7)

### 4.2. Population size (N) effect

Figure 4 shows effect of population size N on infected cases in a Cartesian scale. C, CR and T are fixed and the size N is changed from 3 to 12 and 24 Million. It is deduced that when population size increases epidemic size and period increase, with respective maximums Imax of 25000, 79000 and 128000 reached respectively on days 30, 39 and 42 with respectively epidemic periods of about 41, 56 and 60 days. When N increases the ratio (Imax/N) decreases and respectively given by 0.83%, 0.66% and 0.53%.

**Figure 4.**
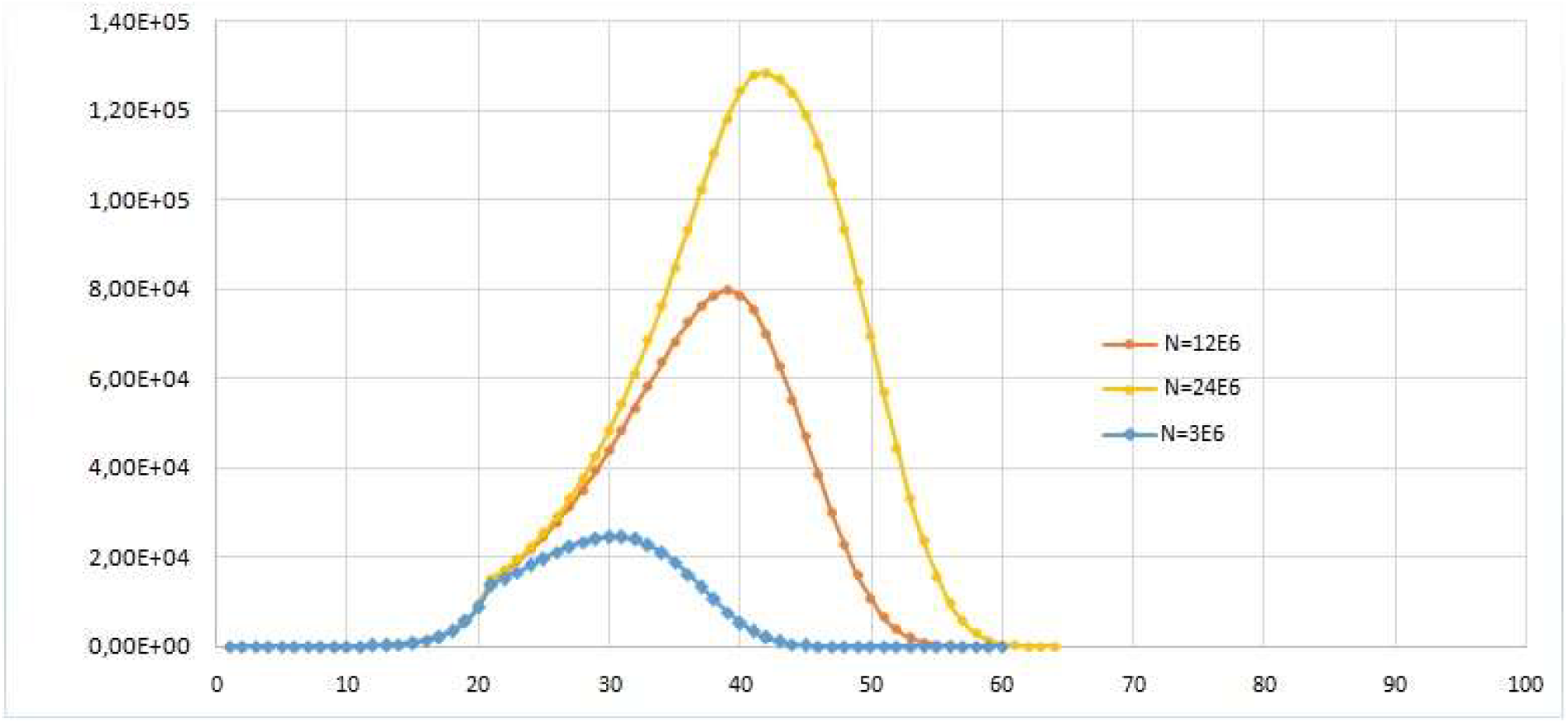
Population size effect on infected cases curve (C=1.62, CR=1.15, T=7)

### 4.3. Tunisian case, population density effect and T effect

Figure 5 presents data in Cartesian scale in Tunisia until Apr 25. On this date, our ministry of health reported 38 deaths and 194 recovered cases. The model is depicted and reproduces actual data tendencies, considering C=1.3 corresponding to initial data fitting in semi logarithmic scale and quarantine and lockdown effects thereafter with CR=1.115. The model reproduces data tendencies with Imax=912 and an epidemic period of 92 days, which means an epidemic spread end at the beginning of June if the same measures are maintained. It is noted that a reduction of C from 1.62 to 1.3 induces a reduction of epidemic peak of about 50 times as deduced when comparing figure 5 with figure 3. Furthermore, it is noted in figure 5 that parameter T when changed from 7 to 25 slightly decreases epidemic.

**Figure 5.**
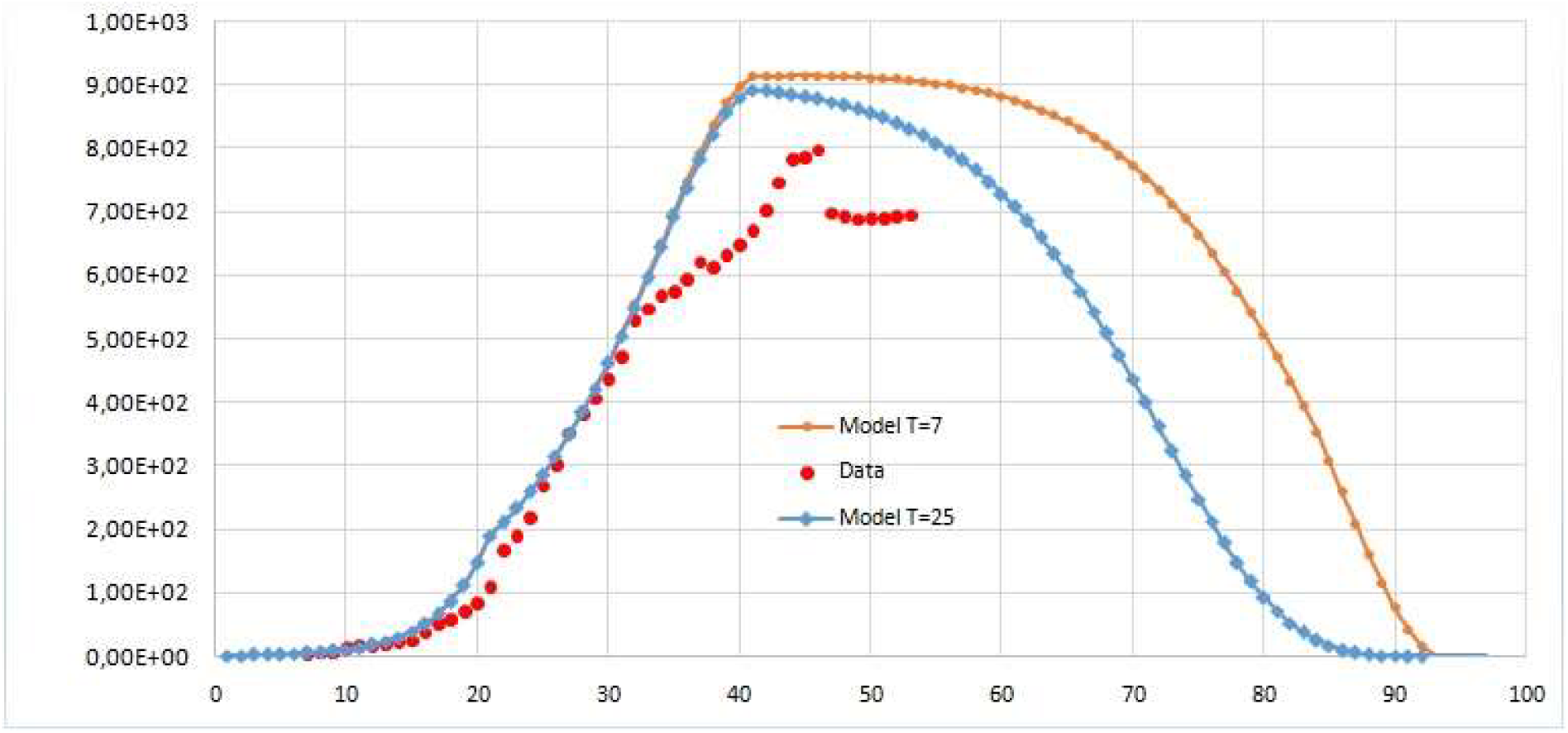
COVID-19 spread in Tunisia (N=12E6, C=1.3, CR=1.115, T=7, T=25)

## 5. Discussion

An evolution law of COVID-19 based on analogy with wave propagation in elastic solids and a damage model is proposed. The key coefficient C is obtained by fitting initial slope of data in a semi-logarithmic scale between day 8 and day 21. Results are very sensitive to this parameter, considered as an intrinsic parameter of population density. For populations of a comparable size, but with coefficients respectively C=1.3 and C=1.62, results show that epidemic size can increase very fast. The second important parameter is CR. It traduces measures like lockdown and quarantine. When controlled, which means decreased, it decreases epidemic size.

## 6. Conclusion

The low predicted epidemic size in Tunisia is essentially due to a low population density (C=1.3) and strict restriction measures (CR=1.115). Population density is inversely proportional to the square of distance d depicted in figure 1 which explains its important effect on transmission.

## Data Availability

National observer for new and emergent diseases, Tunisia

## Author’s Contributions

OL proposed the model and wrote the manuscript.

ML developed medical aspects of the model and wrote the manuscript.

